# Fentanyl dabbing with cannabis retail equipment: an ethnographic and drug-checking study

**DOI:** 10.64898/2026.11.13.26367858

**Authors:** Morgan Godvin, Levi Cecil, Adam Koncsol, Caitlin A. Molina, Megan Grabill, Ruby Romero, Elham Jalayer, Naomi Villanueva, Oscar Arellano, Amanda Cowan, Russell G. Buhr, Honora Englander, Joseph R. Friedman, Philippe Bourgois, Chelsea L. Shover

## Abstract

**Background:** Fentanyl’s entrenchment in U.S. West Coast drug markets brought a shift from injecting to smoking illicit opioids. We characterize the next transition: the rise of ‘dabbing,’ vaporizing fentanyl with equipment sold for cannabis concentrates.

**Methods:** This mixed-methods study combined ethnographic interviewing and participant observation among people who use fentanyl in California and Oregon (2024–2026) with survey data from a Los Angeles drug checking program. After identifying dabbing ethnographically, we added an optional smoking-method item to the survey.

**Results:** Of 108 participants who stated a route of administration, 104 (96%) reported smoking; of 93 who named a smoking method, 81 (87%) reported dabbing. All 11 who first named another method and returned later then reported dabbing. Dabbing was reported at 250 of 405 (62%) encounters with a stated route. We observed participants adapting cannabis equipment. They praised dabbing’s discretion, reduced stigma, and convenience, and its capacity for more large hits, more quickly; the same speed led some to call it wasteful, because it went through their fentanyl faster than other methods. Participants described rapidly rising tolerance and frequent dabbing-related burns, which we also observed. Some who had injected said dabbing approximated its “rush.”

**Conclusions:** Dabbing has become the dominant way to consume fentanyl among people using drug checking services in Los Angeles, and was observed in every other city where we did fieldwork. Clinicians and programs should ask people how they smoke fentanyl, not only whether they smoke or inject; dabbing may reduce some risks while increasing others.

**Highlights:**

- Dabbing has become a dominant way to consume fentanyl in Los Angeles
- Participants repurpose cannabis dab tools and torches for fentanyl vaporization
- Participants described an injection-like rush and fast-rising tolerance as a result
- Dabbing may avert a return to injecting but increases risk of burns and larger hits
- Programs and clinicians should ask how people smoke fentanyl, not only whether

## 1. Introduction

The history of drug use in the United States has unfolded through shifts in dominant substances (Jalal et al., 2018), routes of administration (ROA), and subcultures, each carrying distinct harms (Golub & Johnson, 1999). Until recently, overdose deaths had climbed along a smooth exponential curve since 1979 even as the drugs driving them changed repeatedly (Friedman et al., 2026; Jalal et al., 2018). Generational cohorts often define themselves against the visible harms of preceding practices, as when young New Yorkers who watched crack and heroin injection ravage their elders turned instead to cannabis blunts (Golub & Johnson, 1999). ROA is as much a cultural inheritance as a pharmacological choice; drug trends take hold within generational cohorts and are usually recognized only once well underway (Agar & Reisinger, 2001). The displacement of heroin by illicitly manufactured fentanyl is a recent transition, that ushered in a marked shift from injection to smoking (Ciccarone et al., 2024; Karandinos et al., 2024). Most visible in the western United States but increasingly observed elsewhere, smoking opioids instead of injecting them has been associated with lower all-cause mortality among people in substance use treatment (Karandinos et al., 2025) and fewer skin and soft tissue infections (Megerian et al., 2024), and is expected to reduce other injection-associated harms, including bloodborne infections and endocarditis (Karandinos et al., 2026).

Notably, West Coast heroin markets persisted several years after fentanyl had entered East Coast heroin supplies. When fentanyl arrived in the western U.S. in earnest in 2019–2020 (Shover et al., 2020), it appeared largely as a separate product: counterfeit pills called ‘Blues’ or ‘pressies’ (Godvin & Gonzalez, 2021). As those pills displaced heroin, many people shifted from injecting heroin to smoking fentanyl on foil. The practice of smoking already had precedent in western opioid markets; black tar heroin can be smoked on foil and, being sticky, is not easily snorted (Karandinos et al., 2024; Mars et al., 2016), therefore smoking heroin had long been more common in the West than in the East (Karandinos et al., 2024).

As fentanyl continues to cement its status as the dominant street opioid, novel practices, specialized equipment, and practices of connoisseurship are emerging (Mars et al., 2024). Much of the transition to smoking has come through user-led innovation (Ciccarone et al., 2024; Mars et al., 2024), supplied by equipment for cannabis concentrates, now broadly available thanks to the legal cannabis market. “Dabbing” exemplifies this phenomenon. Borrowed from cannabis concentrate consumption (Loflin & Earleywine, 2014), dabbing transfers this technology to opioid use: a ceramic, glass, quartz, or metal tip, affixed to a tube-like device or inserted in a standard bong, is heated with a butane torch, often allowed to cool briefly, and touched to the substance, and the person inhales the resulting vapor (Figs. 1a and 2). Ciccarone and colleagues (2024) qualitatively characterized dabbing as an emerging ROA among people who use fentanyl in San Francisco, but its prevalence, geographic spread, and everyday practices have not been described.

**Fig. 1.**
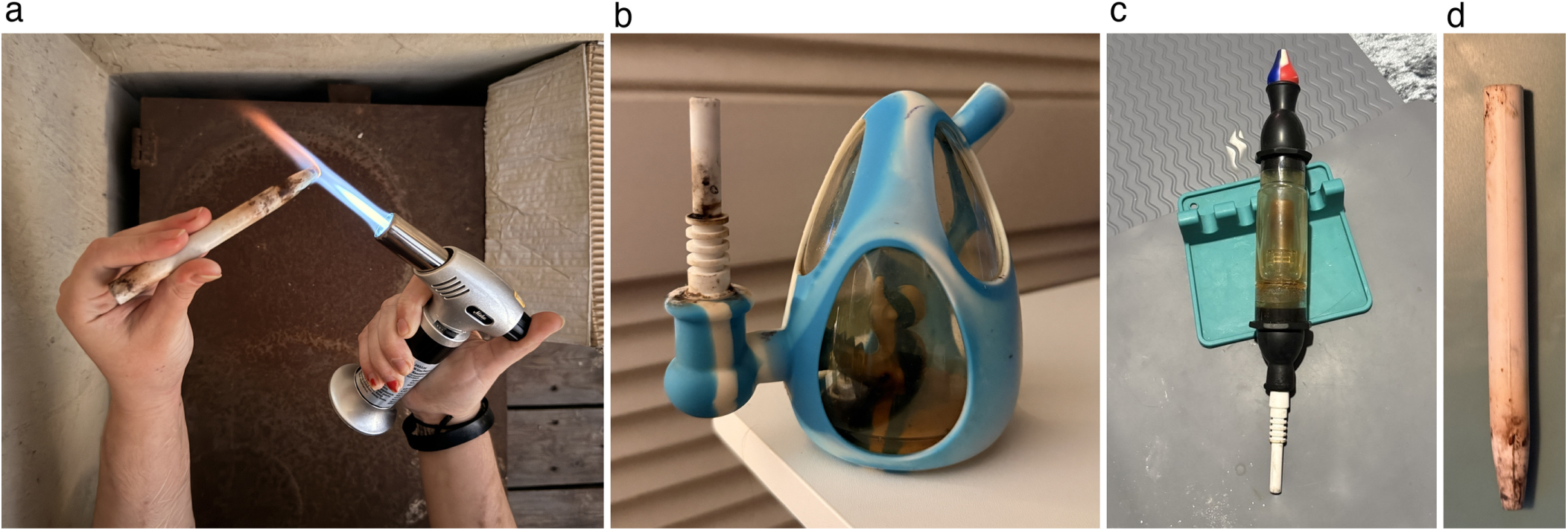
Dabbing in practice. (a) A one-piece ceramic dab straw heated by a participant with a butane torch. (b) A silicone bong fitted with a ceramic dab nail, with residue in its water chamber. (c) A bubbler dab rig with a water chamber, sitting on top of a silicone holder, gray silicone mats are visible underneath it, covering much of the mattress. (d) A one-piece ceramic dab straw with residue at the heated end.

**Fig. 2.**
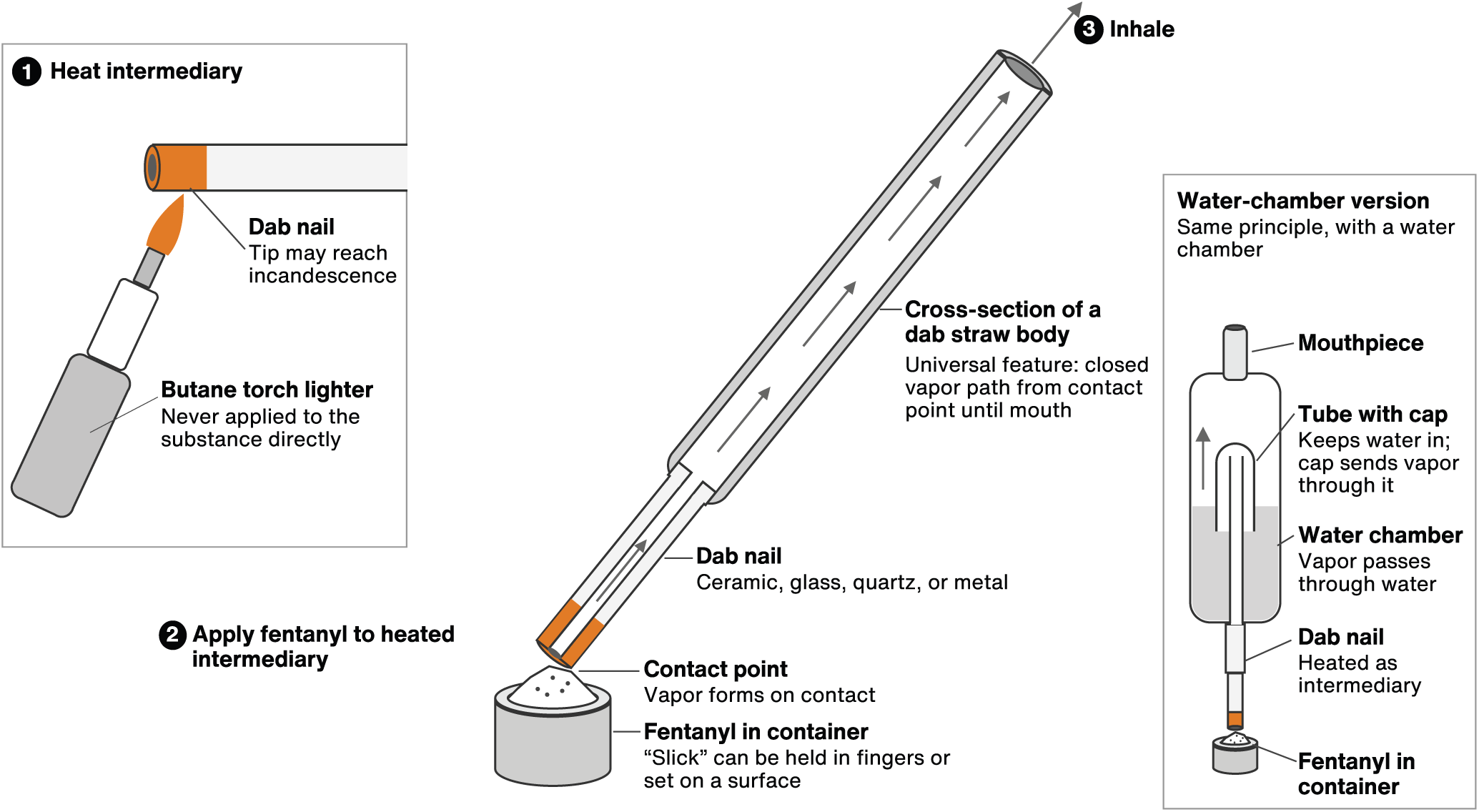
Example diagram of a handheld dab rig body and tip (dab straw/nectar collector). (1) A butane torch lighter heats the dab nail, the intermediary, which may reach incandescence; the flame is never applied to the fentanyl directly. (2) The heated nail is touched to fentanyl in a container (a “slick,” held in the fingers or set on a surface), and vapor forms at the contact point. (3) The vapor travels through the closed path of the dab straw body, shown in cross-section, and is inhaled. The inset shows a water-chamber version that works on the same principle: an internal tube rises above the waterline so water cannot drain into the nail, and a cap over the tube sends the vapor down through the water. Dab nails may be ceramic, quartz, glass, or metal. The diagram describes observed practice, not measured temperatures or dose delivery. Diagram drawn with AI assistance (Claude, Anthropic) from basic shapes; no generative image model was used.

Smoking fentanyl using aluminum foil is virtually identical to the decades-old practice of smoking heroin off foil (“chasing the dragon”) (Hendriks et al., 2001). Dabbing, by contrast, has no direct precedent in earlier waves of illicit opioid use. Both practices are colloquially and clinically termed “smoking,” though both more closely resemble vaporization than combustion (Ondocsin et al., 2025), as does cannabis dabbing (Chen et al., 2025). The person inhales a condensation aerosol, vapor that cools into fine particles off a heated surface (Rabinowitz et al., 2004). Nevertheless, for ease of interpretation, this paper refers to the ROA as “smoking” and the emissions “smoke.”

During participant-observation fieldwork among people who use drugs in Southern California, the ethnographic team (principally the first author MG, with contextual information added by other authors) observed the rapid uptake and normalization of dabbing. We then quantified fentanyl route of administration among participants at a large community-based drug checking program run by the study authors, providing, to our knowledge, the first estimate of dabbing prevalence. Combining these data with clinical expertise in pulmonology and burn injury, we characterize the practice, material culture, social meanings, and potential health implications.

## 2. Methods

### 2.1. Ethnographic methods

This analysis builds on a related ethnographic program on shifting risk environments among people who use drugs in the US–Mexico border region, conducted in Tijuana and other northern Mexican cities beginning in 2018, under separate approvals (Friedman, Bourgois, et al., 2022; Friedman et al., 2023). All ethnographic data analyzed here were collected from 2024 through November 2026, in fieldwork led by MG, under UCLA IRB-21-0901. Observational fieldwork took place in street scenes, encampments, methadone clinics, vehicles, private residences, and, later, smoke shops. MG first observed fentanyl dabbing in Los Angeles in 2024, then continued to observe dabbing through 2026 in Tijuana (Friedman, Bourgois, et al., 2022), San Diego, Portland, and San Francisco. Smoke shops and vape shops across California and Oregon were toured and their inventory photographed.

MG conducted most of the ethnographic fieldwork, spending more than a year in sustained co-presence with people who use fentanyl in California and Oregon, including hundreds of hours observing acquisition, preparation, and consumption. Long-term immersion of this kind reduces social desirability bias and permits comparison of self-reported beliefs and practices with directly observed behavior that cannot be ascertained from epidemiological records alone (Bourgois, 2003; Bourgois & Schonberg, 2009; Friedman, Montero, et al., 2022). All field notes, interviews, and recorded interviews analyzed here were produced by MG; CLS and LC additionally took part in fieldwork and contributed to interpretation as it unfolded. As a community health educator with Drug Checking Los Angeles, LC provided sustained access to community networks, facilitated recruitment for both arms of the study, and, with the drug checking team and community partners, collected the drug checking data. The remaining coauthors contributed clinical expertise, quantitative analysis, or manuscript review and did not generate the ethnographic data analyzed here.

We employed convenience sampling during chance encounters and then snowball sampling as participants introduced us to their wider social networks. We accompanied and informally interviewed participants during routine daily activities. With IRB approval and participant consent, MG audio-recorded 13 conversational interviews in 2025 and 2026 and photographed equipment and practices; dozens more interactions were recorded as field notes. Across fieldwork, MG observed approximately 40 people who use fentanyl and had repeated interactions with 13 key participants, all of whom were interviewed; 9 consented to audio recording (4 were interviewed twice, giving the 13 recorded interviews), and interviews with the other 4 were documented in field notes. Following the photo-ethnographic approach of Bourgois and Schonberg (2009), we treated these photographs as data alongside field notes have reproduced them as figures here. Canva was used to create a diagram to show the mechanics of dabbing.

Analysis was iterative, combining repeated fieldwork with review of field notes and recorded interviews. We developed and refined interpretations through discussions with coauthors, other ethnographers, and community health educators at Drug Checking Los Angeles. We triangulated the perspectives of people who dab, people who smoke on foil, people who inject, and people who sell drugs. Fieldwork continued until we reached saturation on the core practices described below. All names used here are pseudonyms, and identifying details have been altered. To protect confidentiality, accounts from a single individual may appear under more than one pseudonym, and quotes are attributed by name only when continuity requires it. Not everyone engaged in the ethnography participated in drug checking.

#### 2.1.1. Positionality

MG has worked in overdose prevention since 2020, beginning at PrevenCasa in Tijuana, and has administered naloxone to people experiencing overdose since 2011. She has lost people to opioid overdose and is formerly incarcerated, which she discusses publicly, including on social media. MG is a white, working-class woman from the West Coast, and is queer. At six feet tall with a masculine presentation, she was able to relate to men and to feel safe in settings that many women would not enter alone. Her presentation did not fully remove gender dynamics: on a few occasions, the woman in a male-female couple accused her of trying to steal her partner, an explanation that made more sense to them than that MG was earnestly interested in such granular details of drug culture.

MG was occasionally accused of being an undercover police officer. In one instance, the evidence offered was her clean shoes: brand-new, bright white Air Force Ones, a recognized marker of undercover officers trying not to look like police. Not wanting to flaunt a visual marker of her economic privilege, she never wore clean shoes in the field again. In another, a couple suspected her because she could produce drug-checking results and tell them what was in their supply, a capability historically available only to law enforcement. Others, unable to believe that a researcher would take such detailed interest in drug practices, wondered aloud whether she was a police officer, then reasoned that no officer would behave this way, and found the real explanation, that this was academic research, harder to believe still. Because MG is publicly open about her incarceration, participants could verify her account, which usually dispelled the suspicion. That disclosure generated rapport and mutual vulnerability; we treat such enhanced rapport as an ethical issue to be managed rather than a methodological advantage (Harris, 2015).

### 2.2. Drug checking survey methods

Drug Checking Los Angeles operates anonymous, drop-in services at four syringe services programs across Los Angeles County, using portable Fourier-transform infrared (FTIR) spectroscopy, immunoassay test strips, and laboratory-based confirmatory testing. Participants who brought samples for point-of-service analysis completed a brief intake survey. The survey asked what the sample was bought as, what the participant expected it to contain, and whether they had tried it, and recorded an anonymous code, built from partial personal details, that links visits. Staff entered a standard placeholder code when a participant declined to give one, the sample’s origin was unknown, or it was brought on someone else’s behalf; these encounters are excluded from the count of unique participants and cannot be linked across visits.

Participants who brought a sample acquired as fentanyl and had tried it before checking were asked, in an optional item, how they had used that sample; answers describe that sample only, not every route a person uses. A smoking-method question was added later, after changes were observed ethnographically. Participants who reported smoking a sample bought as or believed to be fentanyl could report foil, a bubble pipe/“pookie,” a dabber/nectar collector, or another method. Multiple routes and methods could be endorsed. A recorded route response, including an explicit decline, defined the descriptive cohort; encounters without any route response were excluded. Decline-only responses were excluded from route-prevalence denominators. One response marking both “declined” and “other” was retained as a stated other route. Placeholder codes contributed to encounter-level estimates only.

Participant-level prevalence counts distinct people rather than encounters. if they reported the route or smoking method at any eligible encounter. Route-prevalence denominators include participants or encounters with an actual stated route. Dabbing estimates among smokers retain smoking reports without a named method; estimates specifically among smoking-method respondents include only those who named a method. We separately examined repeated dabbing reports on distinct calendar dates and subsequent dabbing reports among participants whose first answered smoking-method question named another method and who returned on a later date. First answered method does not necessarily mean first service visit or first use of that method. Percentages are rounded to whole numbers, and all denominators are stated explicitly. The quantitative and drug checking methodologies were approved by the University of California, Los Angeles Institutional Review Board (IRB-22-0760 and IRB-22-1273) and some aspects of the work were considered exempt from review as public health surveillance.

### 2.3. Terminology

In this paper, route of administration (ROA) refers to the broad categories recorded in the survey: smoking, injection, insufflation (snorting), and oral ingestion. The ways fentanyl is smoked (dabbing, foil, and bubble pipes) are referred to as smoking methods. This convention follows the structure of the survey (Table 1) and keeps the quantitative and ethnographic findings comparable. It is not a claim that dabbing is simply a variant of smoking; we consider dabbing a discrete, novel route of fentanyl administration (Section 4).

**Table 1.** Reported fentanyl routes and smoking methods among unique participants and sample encounters, Los Angeles County, July 2024–November 2026.

| **Route or smoking method** | **Unique respondents, n** | **Sample encounters, n** |
|---|---|---|
| **Smoking** | 104 | 392 |
| Dabbing | 81 | 250 |
| Foil | 33 | 122 |
| Pookie / oil burner / glass bubble pipe | 27 | 120 |
| Other identifiable smoking method | 0 | 0 |
| Smoking method unspecified at ≥1 encounter | 14 | 19 |
| **Injection** | 8 | 14 |
| **Oral ingestion** | 3 | 5 |
| **Insufflation (snorting)** | 2 | 2 |
| **Other route** | 3 | 4 |
| **Declined without stating a route** | 15 | 34 |

Note. People are counted once per route or method if they reported it at any eligible encounter; categories overlap. The cohort comprises 438 encounters, including 381 linked to 116 participants and 57 with placeholder codes. Actual routes were stated at 405 encounters and by 108 participants. Fifteen people declined to state a route at least once; eight never stated one. Fourteen smokers had at least one encounter with an unspecified method, and 11 never named a method. One “declined,other” response is counted as a stated other route. Placeholder encounters contribute to encounter counts only.

There is no accepted taxonomy of the ways fentanyl is smoked. This paper uses “dabbing” as the umbrella term for heating fentanyl via an intermediary device and inhaling the vapor. Smoke shops sell the devices under several names, including dabbers, dab straws, and nectar collectors; people who use fentanyl used these interchangeably. A “dabber” can refer to the device, alternatively called a “dab rig,” yet a “dabber” can also mean “someone who dabs.”.Tips refer to the interchangeable pieces meant to be heated that are inserted into a dab straw, though with one-piece glass or ceramic dabbers, it reverts to the plain English definition of the very end of the device, which is still the portion that is heated. Smoking through a bong or bubbler is classed here as dabbing because their use for fentanyl requires a dabbing-specific tip — a dab nail, “banger” (glass cup-shaped bong insert), or similar attachment meant to be heated directly first, to which the fentanyl is then applied at a separate, later step (Figs. 1b–c and 3).

**Fig. 3.**
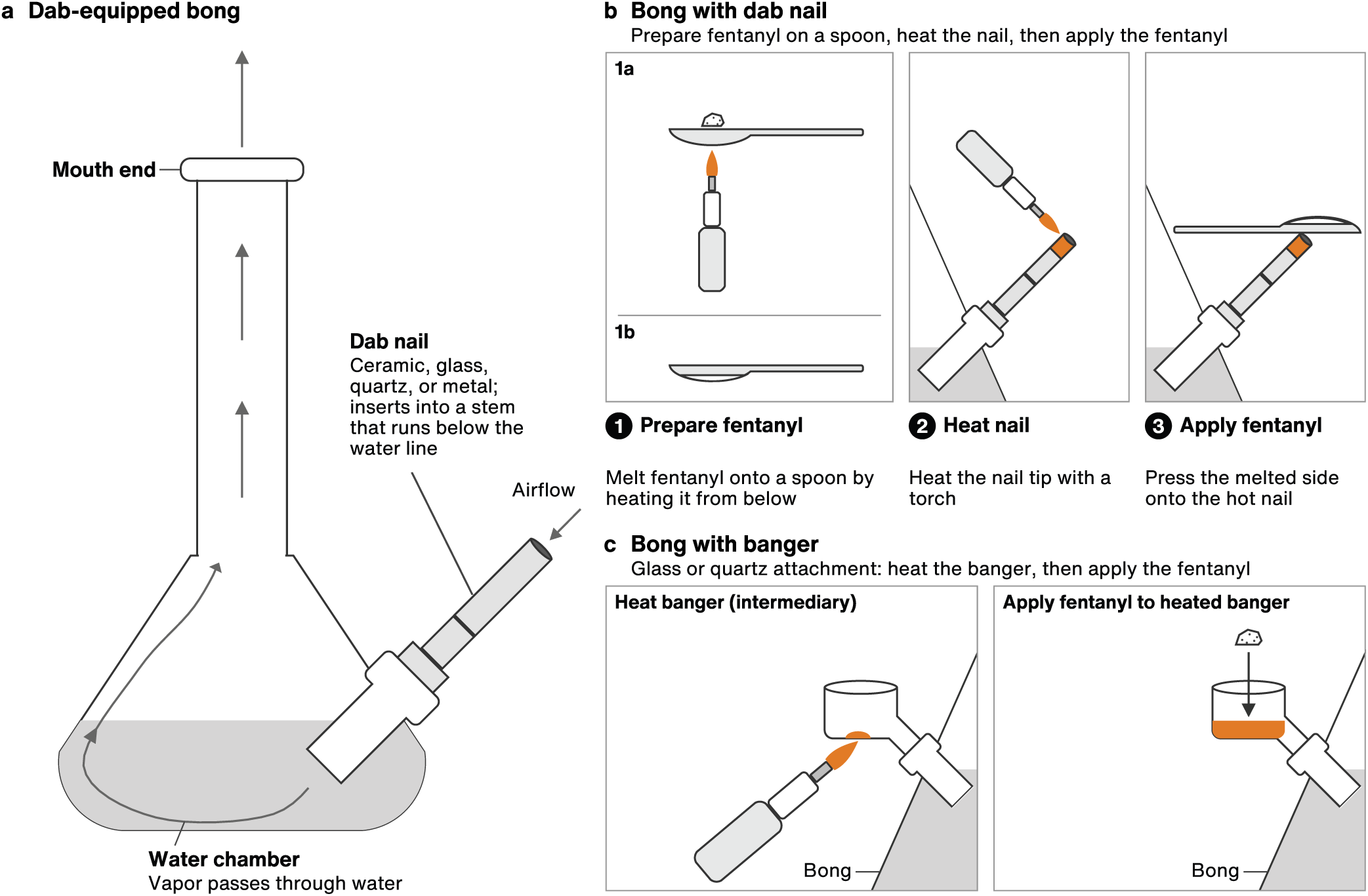
Dab-mounted bongs and other attachments. (a) A dab-equipped bong. The dab nail (ceramic, glass, quartz, or metal) inserts into a stem that runs below the water line; vapor passes through the water chamber to the mouth end. (b) Bong with a dab nail: fentanyl is melted onto a spoon heated from below (1a, 1b), the nail tip is heated with a torch (2), and the melted side of the spoon is pressed onto the hot nail (3). (c) Bong with a banger: the glass or quartz banger is heated first, and fentanyl is then applied to the heated banger. The diagram describes observed practice, not measured temperatures or dose delivery. Diagram drawn with AI assistance (Claude, Anthropic) from basic shapes; no generative image model was used.

Consequently, free-text “other method” answers naming a bong, vape, or crack pipe were coded as dabbing because an intermediary is heated, then applied to the fentanyl. When crack pipes are used to consume fentanyl, the only feasible way to do so is to use them as dab straws. Heating fentanyl with an open flame, as done for crack, would incinerate it before the vapor could be consumed.

We did not consider “oil burner” bubble pipes to be dabbing, because the fentanyl itself is heated with flame, albeit through the barrier of the glass bubble. Hammer pipes were not observed, but would represent a gray area, as they can be used in either way. In this text, when referring to pipes, we mean the ones sold under the euphemism “oil burner,” bubble-style glass pipes known colloquially as “pookies.”

We distinguish a hit, a single inhalation, from a dose, the amount of fentanyl a person means to consume in one session, which may take one hit or many; with injection, the two are one and the same. The dose is what is in the syringe. With smoking, they become distinct, though sometimes overlapping, concepts. How much of a dose actually reaches the person depends on both technique and equipment.

## 3. Results

### 3.1. Quantitative results

Between July 2024 and November 2026, 438 fentanyl-sample encounters had a recorded route response: 381 linked to 116 unique participants and 57 recorded under placeholder codes. Among 108 participants who reported a route of administration, 104 (96%) reported smoking. Of these, 93 reported a smoking method at least once, and 81 of those 93 (87%) reported dabbing (Tables 1 and 2). Dabbing was reported by 81 of 108 participants with a stated route (75%) and 81 of 104 who reported smoking (78%).

**Table 2.** Dabbing was reported by 81 of 93 smoking-method respondents (87%): prevalence and subsequent reporting under explicitly defined participant and encounter denominators.

| **Unit / finding** | **Population forming the denominator** | **n/N** | **%** |
|---|---|---|---|
| People: smoking | Stated an actual route | 104/108 | 96 |
| People: dabbing at least once | Reported a smoking method at least once | 81/93 | 87 |
| People: dabbing at least once | Reported smoking (with or without a method) | 81/104 | 78 |
| People: dabbing at least once | Stated an actual route | 81/108 | 75 |
| People: named another method first, then dabbed on a later date | Initially named a smoking method other than dabbing; returned on a later date | 11/11 | 100 |
| People: dabbing on multiple dates | Named a smoking method on at least two dates | 25/33 | 76 |
| Encounters: smoking | Stated an actual route | 392/405 | 97 |
| Encounters: dabbing | Stated an actual route | 250/405 | 62 |
| Smoking encounters: dabbing | Reported smoking, including method nonresponse | 250/392 | 64 |
| Smoking encounters: dabbing | Recorded a smoking method | 250/373 | 67 |

Note. Participant estimates count each person once. Encounter estimates may include repeated responses from the same person. Smoking-method categories overlap. Decline-only route responses are excluded from actual-route denominators. Smoking encounters without a method answer remain in the all-smoker denominator. The 11/11 comparison begins with each participant’s first answered smoking-method question. All 33 participants with method answers on at least two dates reported dabbing at least once; 25 reported it on multiple dates. A later report does not establish when dabbing began.

Eleven participants whose first answered smoking-method question named a method other than dabbing returned on a later date, and all 11 subsequently reported dabbing. Among 33 participants who reported a smoking method on at least two distinct dates, all reported dabbing at least once, and 25 (76%) reported dabbing on multiple dates.

Across 405 fentanyl-sample encounters with a reported route of administration, respondents reported smoking at 392 (97%) and dabbing at 250 (62%). Dabbing was reported at 250 of 392 smoking encounters (64%), or 250 of 373 smoking encounters with a recorded smoking method (67%). Foil was reported at 122 smoking encounters (31%) and a pipe at 120 (31%); methods could overlap. Nineteen smoking encounters lacked a recorded method. Dabbing was the sole route and smoking method reported at 150 of the 405 encounters (37%).

Injection was reported by 8 of 108 participants with a stated route (7%); 5 of those 8 (63%) also reported dabbing at least once. At the encounter level, injection was reported at 14 of 405 encounters with a stated route (3%). Nine of those 14 injection encounters (64%) also included dabbing for the same sample; four reported injection as the sole route.

### 3.2. Ethnographic results

#### 3.2.1. From foil to dabs: the emergence of a practice

Foil was the dominant fentanyl smoking method in earlier fieldwork and remained commonplace when MG resumed fieldwork in Los Angeles in 2024 and first observed dabbing there; when and where people who use fentanyl first began dabbing is unknown.

Participants typically encountered dabbing through demonstration within their networks, often by sellers. Jack, a Southern California resident in his 30s, first saw it when he went to buy a gram from a dealer advertising openly on Craigslist:

“He handed me this chunk that looked like plastic — it had this melted plastic yellow sheen on it. And I was like, ‘Bro, what is this?’ And he was like, ‘Chill out, I just took a dab hit off it.’ And I was like, ‘You did a what?’…He torched up the nail and let me take a hit off of it. And I was like, holy shit.”

Adoption was not instantaneous. Jack described ‘a psychological affinity to foil’ built from repetition; his first solo attempt with a glass nectar collector and $4 torch lighter produced ‘such little smoke’ that he returned to foil. The same seller later handed him a full-sized torch: ‘As soon as I started using that blue torch he gave me, I understood.’ With sufficient heat, the practice transformed: ‘I blew out this giant plume of white smoke…it was like five or ten hits from foil in one pass.’ It was ‘life-changing…I never went back.’

Dabbing spread through self-teaching and increasingly through shared knowledge:

“I didn’t know very many other people that were doing it, so I just had a lot of trial and error, buying stuff at smoke shops — how do you deal with it when your nectar clogs? If you run out of dope, how do you scrape it?…I figured all that stuff out and just kept iterating. And as time went on, I saw more and more people dabbing…I’d start to recognize the signs. When I met other friends who also dabbed, we were swapping this knowledge back and forth.”

Other trajectories ran through the exhaustion of alternatives. Alex, who had injected for years, switched to foil when his ‘veins gave out’ — but hated it, and kept attempting to inject anyway. Once someone showed him how to dab, he never returned to either. His partner Jane was blunter about foil:

“Oh my god, no. I hated foil. Even at the time I hated foil.” — Jane, asked whether she would ever go back to foil; field note, May 2026

Both Alex and Jane, who had moved from the East Coast, had injected daily, and both said they had not injected at all in the six months since they began dabbing. For Jack, dabbing had eliminated injection’s rationale: “You got to find a vein, you got to have syringes, you got to poke yourself…when you could just take a fat dab rip. So what is the value add to injection?”

Across the fieldwork period, nearly all individuals in the network who tried dabbing with a full-sized torch went on to adopt it as their preferred smoking method, although some survey participants reported more than one smoking method. Most of those still using foil had either not tried dabbing or had tried it only with a cheap torch lighter, as Jack first did. For some, dabbing’s startup cost was off-putting, and people who did not dab tended to prefer bubble pipes, sold three for $5 across Los Angeles and given out by syringe services programs, though they rarely lasted more than a few days. Dabbers can last for months, but butane is a recurring expense.

Among people encountered during fieldwork, most preferred dabbing to pipes, and pipes to foil, while accessibility ran in the opposite direction: foil was cheapest and most readily available, followed by pipes, and dabbing required a larger initial investment in equipment. Taking up dabbing meant adding it to a repertoire, not abandoning other methods; smoking method was fluid and contextual. Because dabbing goes through fentanyl faster (Sections 3.2.3 and 3.2.5), even people who preferred it rotated methods as money allowed, dabbing when flush and switching to an oil burner to conserve. In the survey, two in five participants who reported dabbing (34 of 81, 42%) also reported another smoking method: foil (28, 35%), an oil burner (20, 25%), or both. Among participants who answered the smoking-method question on at least two distinct dates, 21 of 33 (64%) reported a different combination of methods on different dates, whereas only 4 of 37 (11%) who answered the route question on at least two dates reported a different route or combination of routes across their visits.

#### 3.2.2. Artisanal assembly and the material culture of dab rigs

We observed elaborate customization among people who dab, including dab rigs made from bongs, bubblers, and custom-built apparatuses (Fig. 1b–d). Devices are most often assembled from a body and a tip, each in a range of materials, shapes, and sizes (Fig. 2). Tips are ceramic, glass, quartz, or metal and are heated directly; bodies are glass, silicone, or combinations thereof, which interrupt conduction and remain cool enough to handle. One-piece ceramic dab straws were also used (Fig. 1a, d), and crack pipes given out as safer smoking equipment were repurposed as dab straws.

One-piece glass dabbers, distributed by at least two overdose-prevention organizations in the region, were not considered ideal because fentanyl’s sedation can cause people to drop things, and a dropped glass dabber shatters: “You take a fat rip, it kicks in, and you nod out and drop your dabber on the ground,” one participant explained. “That’s going to happen a million times a day.” Glass also cools faster than other materials, so less drug can be consumed per hit. Silicone bodies were more commonly observed, including small silicone bongs. A glass dabberstylized as a black bear is displayed alongside silicone devices and firearm-shaped dab straws and torches illustrate this range of materials and novelty forms at retail (Fig. 4a–b) Glass cannot stretch to grip a tip the way silicone does, so glass tips are held in glass nectar collectors with plastic clips (Fig. 4c). Many participants preferred ceramic tips, describing them as staying hotter longer. Although many store-bought devices ship with metal tips, these were rarely observed in use: “The metal ones taste bad…It’s cheap imported [metal]. I don’t trust this metal. I don’t want to superheat it and then inhale it.” An Amazon listing for ceramic tips used both euphemistic sandblasting terminology and the insider slang of “nectar collector” in its product title, unable to sell cannabis dabbing equipment openly due to Amazon’s terms of service, which didn’t stop several participants from purchasing their equipment from the site.

**Fig. 4.**
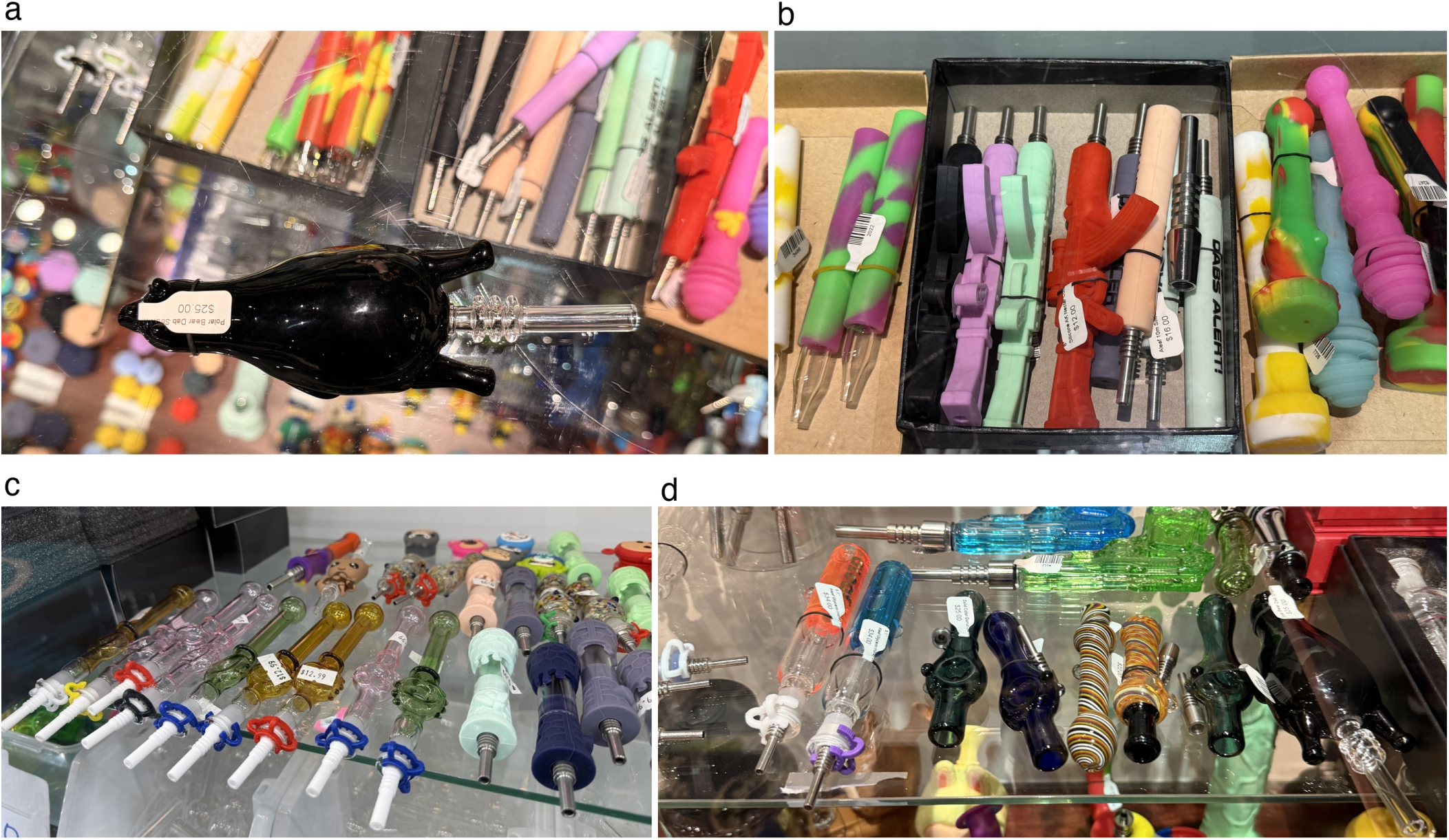
Dabbing equipment at retail. (a) A glass dab rig stylized as a black bear ($25), in front of silicone dab straws. (b) Novelty-shaped silicone nectar collectors with metal tips, $12–$16. (c) Glass nectar collectors with ceramic tips held on by plastic clips, beside silicone collectors with metal tips. (d) Novelty glass dab rigs above silicone water pipes.

Torches cost from a few dollars to approximately $50; tips, dab straws, nectar collectors, and assembled devices $5–35; and electronic battery-heated dab rigs $30–$300 or more. Prices were posted at the smoke shops toured during fieldwork and on Amazon and other online retailers, 2024–2026. Torch lighters at retail are shown in Fig. 5. Devices bought assembled are often customized further — the stock tip swapped out, or built from store-bought tips inserted into repurposed mouthpieces, such as the red rubber mouthpieces syringe services programs hand out to reduce disease transmission from pipe sharing. Equipment cost also shaped method choice: “If you just spent your last 20 bucks on a bag, you’re not going to want to spend” on equipment, whereas foil “you can pretty much get for free, anywhere, anytime”; several participants framed even their occasional continued use of foil as something poverty imposed on them, not something they preferred. Butane became more widely available over the study period and is now a common sight at corner stores and even chain convenience stores, e.g. “7-11”. Empty butane canisters have become part of the milieu of urban detritus, though not nearly as common as crunched-up burnt foils had once been.

**Fig. 5.**
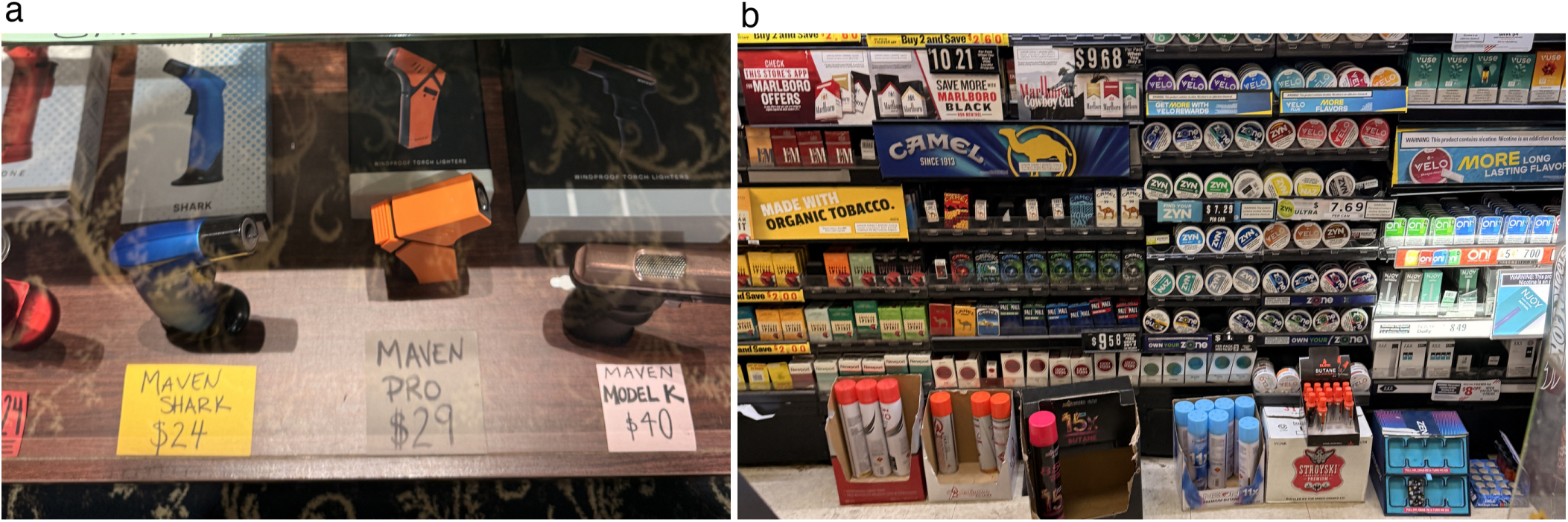
Torches and butane at retail. (a) Torch lighters priced $24–$40 at a smoke shop. (b) Butane refills and small torch lighters at a corner-store counter.

Electronic dab rigs, vaporizers, and e-nails were seen more often in recent fieldwork, but, being expensive, only among people with more access to resources. Street fentanyl is typically cut with bulking agents and other additives, with a mean fentanyl purity of 12.5% in our previously published study; therefore, 87.5% by mass of product sold as fentanyl is actually bulking agents and other compounds (Shover, Koncsol, et al., 2025; Godvin et al., 2026). Those impurities clogged devices designed for cannabis concentrates; one participant would not risk returning a device to the store due to the incriminating residue, though it left him out $200 and broke within hours of purchase.

Fentanyl is usually held and dabbed from a small silicone container (a “slick,” a term borrowed from cannabis dabbing), which people held in their fingers and brought up to the heated tip to shield their fingers from burns, like an inverted thimble; the slick also catches any fentanyl that melts or breaks off and, sold with a lid, doubles as a sealable container that does not look incriminating. Some people skipped the slick and held the chunk in their fingertips, which caused mild burns when the hot tip grazed an adjacent finger (Section 3.2.6). Others brought the fentanyl to the heated element instead, dumping it onto the hot surface or melting it onto a metal spoon that is then turned over and pressed down onto a dab nail sticking out of a bong (Figs. 3b and 6a).

**Fig. 6.**
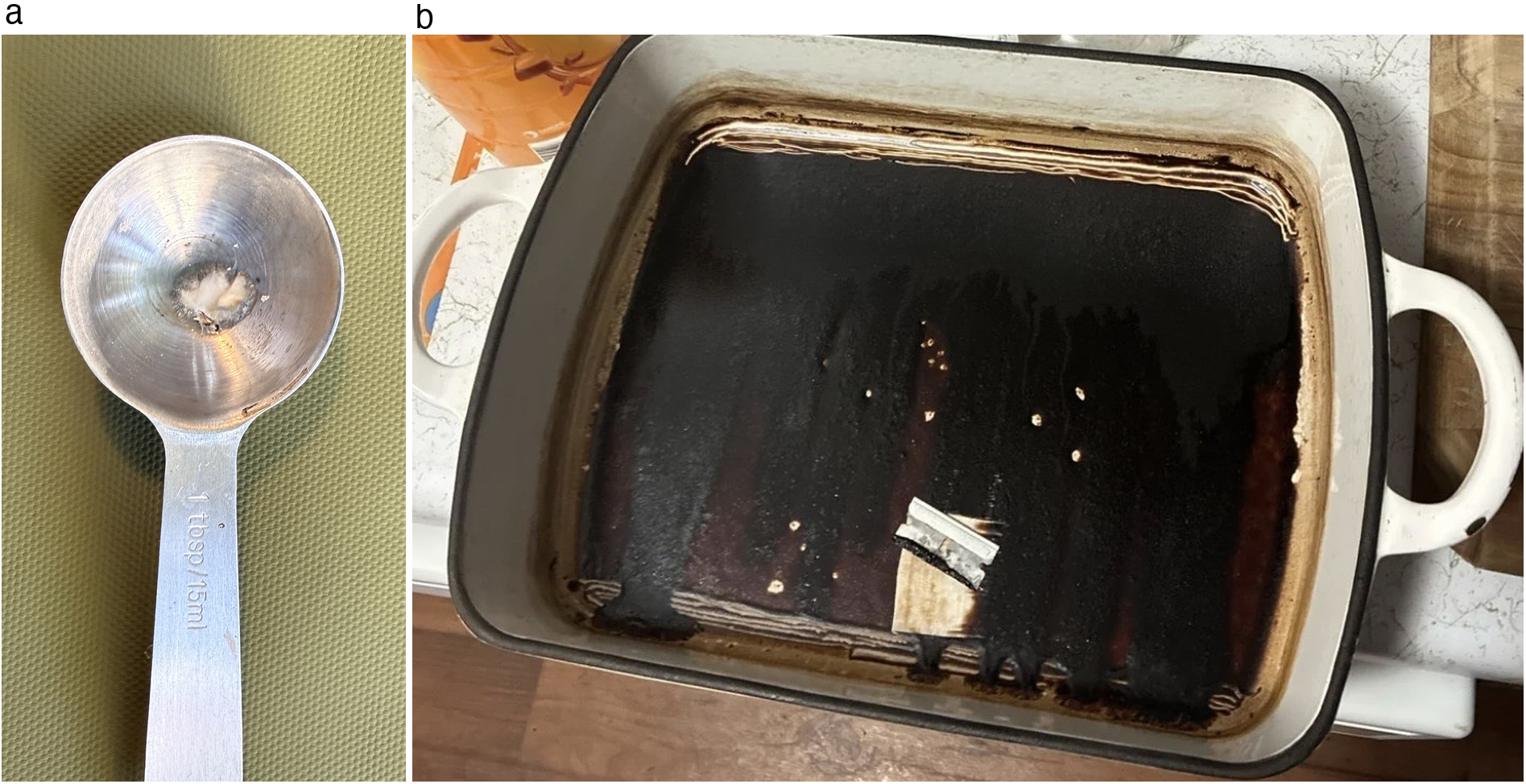
Melted fentanyl and residue. (a) A metal tablespoon with melted fentanyl in the bowl. (b) Fentanyl residue (“reclaim”) left in a baking dish after bong water was evaporated, with a razor blade used to scrape it up.

Retail supply has expanded in step with demand, and participants experienced the change as a rapid cultural shift, even as foil persisted (Section 3.1):

“Dabbers are infinitely customizable. All my friends are dabbing [now]…they’ll go from not knowing how to dab to, the next time I go over there, they have these wicked custom-built rigs. People are modifying bongs, melting a gram or two into a tablespoon and then touching the tablespoon to the heated dab nail, giant three-footers, just DIY — and everybody’s got their preferences now…It’s generated this insane cultural shift.” — Jack

Alex owned at least eight customized rigs. His eyes lit with pride as he showed visitors his latest “Frankenstein” build.

This artisanal diversity, sustained by continual peer exchange of parts and know-how, constitutes a visible, durable material culture among people who dab fentanyl.

#### 3.2.3. Technique, hit size, and titration

Participants typically heated the tip with the open flame of a butane torch for 10–30 seconds and often let it cool, if only briefly, before touching it to the fentanyl. An ordinary cigarette lighter cannot heat ceramic sufficiently over a wide enough area. Flames range from under half an inch to nearly a foot; larger flames heat tips faster but burn more butane, cause more accidental burns, and are less portable. They also produce more noise, and discretion has two competing needs, speed and quiet. A quiet torch has a small flame that struggles to heat a 14 mm nail, and participants weighed these factors against one another. The decisive variable is reaching adequate temperature: as one participant put it, “You really had to get it super white hot.” A water-pipe setup with a ceramic dab nail is shown in Fig. 1b.

Participants described longer inhalations and more concentrated smoke using dabbing than with foil. People who bought several days’ worth of fentanyl supply at a time would often dab off one large chunk. The largest observed being dabbed off during fieldwork was a twenty five-gram rock. How much each draw consumed was hard to judge. With the tip against the fentanyl, nobody can see how much is vaporizing, and the only running measure is the shrinking chunk, sometimes smoked off for days or weeks at a stretch (Fig. 7). One participant called it “a blunt force instrument”: “You can’t exactly titrate your dose like you can on foil…But holy hell, can you get some fat rips off it.”

**Fig. 7.**
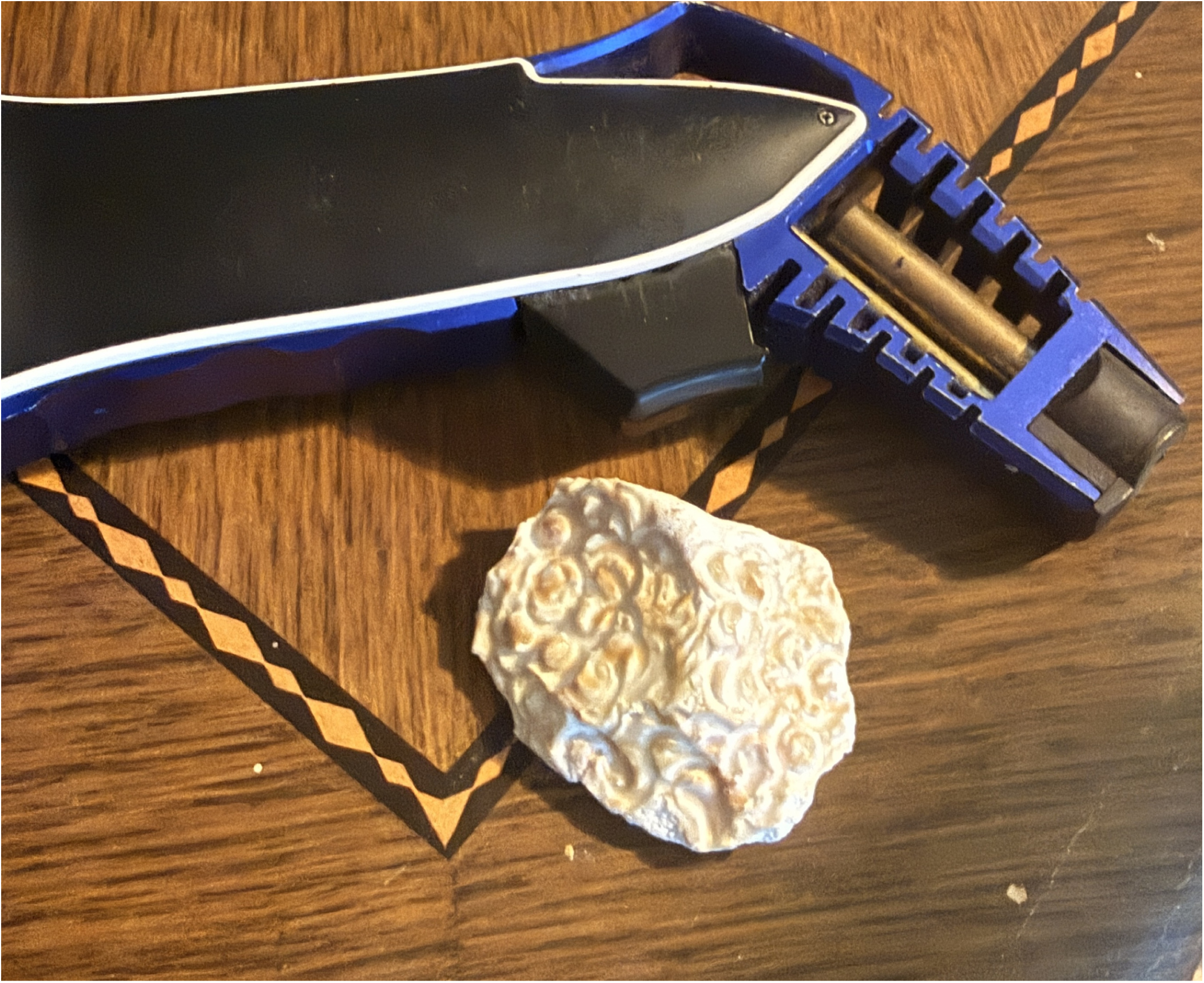
A large fentanyl chunk beside the blue torch lighter a seller gave Jack in 2024 (Section 3.2.1).

The large inhaled hit was experienced as replicating the rush of injection (Jack: “it almost reminded me of shooting up…you took in such a large amount that you would immediately feel it going through your body”), echoing accounts from San Francisco (Ciccarone et al., 2024). Taylor estimated one large hit at 100–200 milligrams of total drug product (“a point, maybe two points”), while acknowledging the unknowns: “How much of that is getting trapped in the dabber and how much is actually converting to smoke that you inhale? I don’t know.” Because batches varied in how “harsh” they were, the feel of smoke in the lungs was an unreliable guide to how much had been taken.

Dabbing can also be done quickly, permitting large hits taken in quick succession: “you could smoke a half gram in a couple hits, just in a few minutes.” Speed cut both ways: it returned hours of the day once lost to foil (“rip, rip, rip, go on about your life”), but with “not really a satiation point,” participants described tolerance climbing fast, “up to doing many grams a day that way.” As one participant put it, “Dabbing is a monster…it’s a double-edged sword, because then your tolerance goes up so much, so fast.”

Participants had strategies for modulating the size of each hit: tips with less surface area, tips made of something other than ceramic, less fentanyl in the slick, and touching less of it with the heated tip; some made only the briefest taps with the very corner of the nail to conserve. Heat management was a trade-off: “You can’t get it too hot, because then you’re degrading the fentanyl. But if you don’t get it too hot, then the device cools so quickly you can’t take a big draw.” Heating times varied with tip material and torch strength, so participants had no way to know a tip’s temperature. Visible red heat was treated as the only reliable sign that it was hot enough, and many also understood it to be too hot: Alex and Jane both heated the ceramic to incandescence and then let it cool for several seconds before touching it to the fentanyl, because at full heat it “burns up the fentanyl.” Styles ranged from precise to maximal. Alex favored small, precise hits at medium heat. Jane heated the ceramic nail of a plain silicone dab straw until it began to glow red, set the torch down, picked up the slick holding her chunk, put the other end of the straw in her mouth, and brought the fentanyl up to the tip, gripping the slick between thumb and index finger. The glow had mostly subsided in the few seconds it took to make contact. She inhaled until her lungs were full, tapping the nail rhythmically against the rock as wisps of smoke escaped from the sides.

Whereas people once portioned a small dose of fentanyl onto foil, people who dab often smoke off a large chunk, however it came from the seller. The form of the supply interacts with the method: loose powder is hard to dab, because it can be drawn through the straw before it vaporizes, leaving a mouthful of bitter powder. Swallowed fentanyl has substantial first-pass metabolism, with roughly 30% bioavailability and a delayed peak (Lötsch et al., 2013; Streisand et al., 1991); people counted powder in the mouth as wasted, although a swallowed dose could add to smoked doses after a delay. This has created a market preference for fentanyl “rocks” or “chunks” (Fig. 8a), which are also seen as less likely to have been adulterated. When loose powder circulated, participants pressed it back into chunks, bound it together with water, carefully melted it onto the slick (sometimes singeing the silicone), or heated it on a metal spoon from below, which reminded one participant of the days when every spoon in their home was bent and charred from preparing heroin injections.

**Fig. 8.**
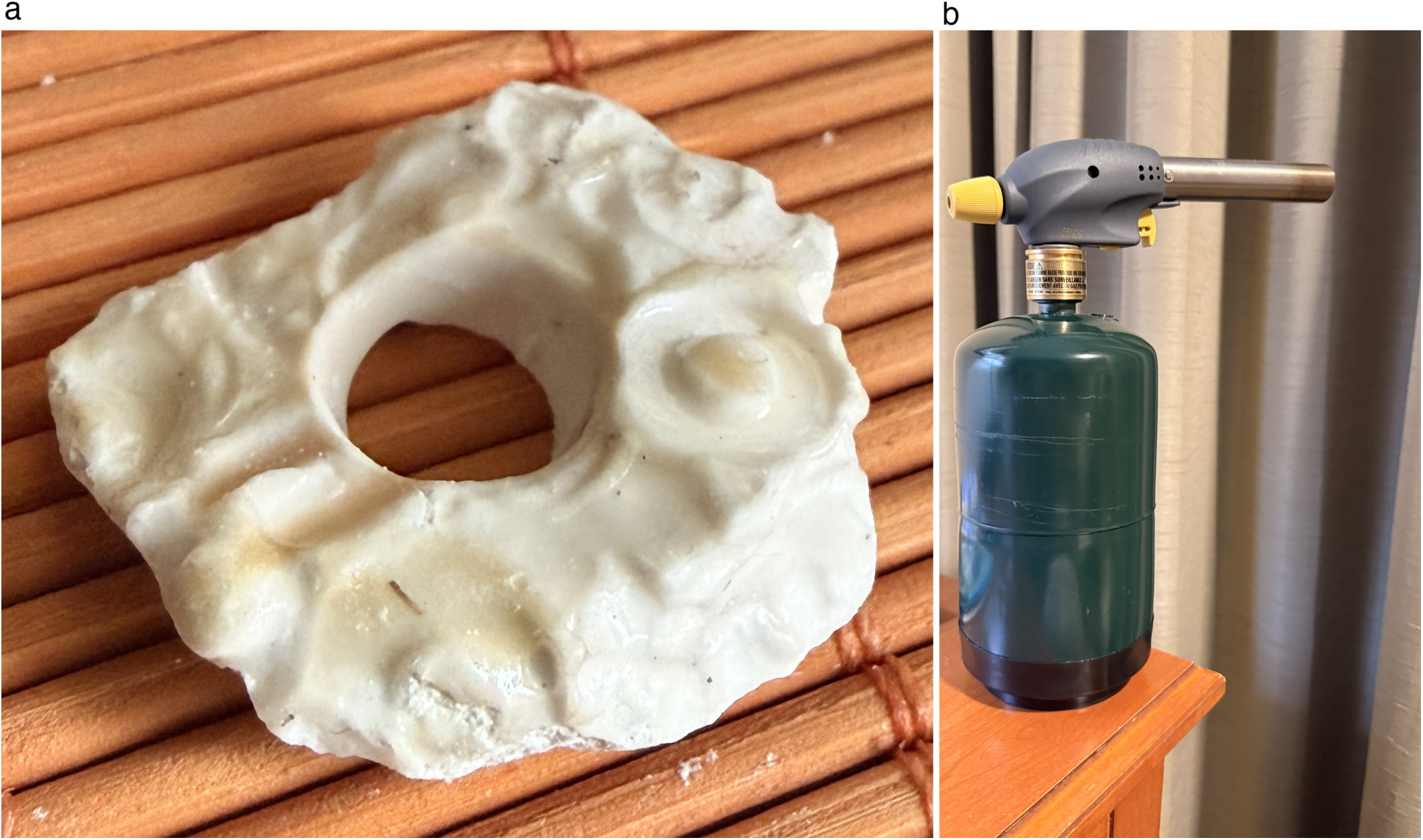
Melted fentanyl and a blowtorch. (a) A fentanyl chunk with a glossy, melted surface surrounding an opening. (b) A blowtorch head mounted directly on a fuel canister; one participant heated the nail of a 3-foot bong with a comparable setup (Section 3.2.5).

#### 3.2.4. Discretion, mobility, and stigma management

Participants generally valued dabbing’s discretion and convenience outdoors. Torch flames stood up to wind, and the closed path kept smoke from blowing away before it could be inhaled, unlike foil, which could require shielding both lighter and smoke from the wind by conspicuously covering one’s head with a towel or blanket. As one participant put it, “With dabbing there’s none of that. I can sit outside in wind, torch up my nail — the torch is such high pressure it stays on even through windy conditions — and then you touch the nail directly to your stuff, so there’s not a gap of air where wind could get in.”

Dabbing is generally less conspicuous than smoking off foil and can be done in a wider range of settings. “You can just be sitting there all nonchalant, fire up the torch, and hit the dab nail sitting on your lap without your head bent over…really quick, put it up to your mouth, tap, tap, tap, and you’re done,” one participant explained. “That’s protective of getting the cops called on you, or getting busted.” Several described foil as the defining public image of the current era: slow, legible to police and passersby, and, because sessions stretch across hours, lending the appearance that drug use had “skyrocketed” even if it had not.

Buying dabbing equipment, ostensibly made for a legal substance, drew less stigma from store employees than buying syringes or glass pipes: “People knew that those little glass roses were actually crack pipes, and that an oil burner is actually a meth pipe. But with dabs, people actually think you’re dabbing weed…You don’t feel as dirty when you go to the store to buy it.” Because smoke-shop staff assumed customers were smoking cannabis, Sam could discuss tools and techniques openly, which he contrasted with injection expertise, long “locked away, relegated to the shadows.” If a family member or police officer discovered their equipment, participants could lean on the social acceptability of cannabis to explain it away without disclosing fentanyl use, which remains heavily criminalized.

Dabbing’s discretion also sustained employment. Several participants in this network held regular jobs and used fentanyl several times a day. Fentanyl’s short duration of action imposes a redosing rhythm that prior opioid eras did not: as one participant put it, “You could go a whole shift of work without doing a shot of heroin, but fetty is a different beast”; these participants smoked at least every few hours. Some used the bathroom; others dabbed in their cars at work, with strategies to further mitigate risk:

“Just bring a [nicotine] vape to your mouth when you blow out the smoke.” — Jane, on dabbing in the car; field note

Chris taped his dabber to a hookah hose, making a seal, so he could dab while driving long distances: he could hold his dabber in one hand, the steering wheel in the other, and the hookah mouthpiece in his mouth, driving down the road without making movements suspicious to other drivers. Participants acknowledged the risk of dabbing in public and said they did it as little as possible: “but what’s the alternative? I have to go to work, you know. I do what I gotta do.”

#### 3.2.5. Folk pharmacology and social meanings

Participants articulated a folk pharmacology in which dabbing recovers a sensation fentanyl otherwise withholds. Sam highlighted dabbing’s ability to restore a perceptible “rush”: “Fentanyl is kind of a shit drug. It’s not euphoric, it’s not warm and fuzzy. Half the time you can’t even perceive that you’re high. All the time you see people nodding out — you’re like, ‘Bro, you should sit down,’ and they’ll be like, ‘What? I’m not even high,’ and then nod out again. They’re not lying…It’s a lights-out drug — there is nothing between zero and nine, then ten you’re asleep. But dabbing was able to recreate [the rush] because of the rapid onset of such a high dose. You could actually feel something.”

Sam was not alone. Participants in this network lamented the loss of heroin’s long, warm euphoria, an hours-long reminder that you had consumed opioids. Opinions diverged on whether fentanyl was a lost cause entirely, or whether, in the right circumstances, it could still deliver the fleeting “rush” once thought possible only by injecting. No one said that dabbing fentanyl created a rush like heroin. Several said that a big enough dab hit absolutely produced a rush like injecting, and that belief kept some people innovating, perpetually chasing bigger hits.

Corey showed where the chase leads. In May 2024 we watched him sitting on his bed, casually heating a ceramic nail in a 3-foot bong with a full-sized blowtorch attached to a fuel canister (a comparable setup is shown in Fig. 8b). He tapped a several-gram chunk to the nail, inhaled, and slumped down the wall. We feared an overdose until he exhaled a thick cloud and let out a “now that’s what I’m talking about,” his voice a lower octave than it had been just moments before. A fat enough hit, he said, gave a rush almost like injecting. He had started on a regular dab straw and built bigger setups as tolerance grew, which took money, both to buy supplies and to buy enough fentanyl to have a tolerance that high in the first place. His cycle was common: people impressed by a rig’s capacity soon found its biggest hit too small as tolerance caught up with each upgrade. Many concluded that a single hit has a ceiling and simply took more; we watched people take up to a dozen large hits and remain awake and alert. Smoke wisped from the bong; big hits meant waste.

Waste itself was contested. The word did double duty. It named material loss: fentanyl drifting off as smoke, degrading on an overheated nail, or left clinging inside the dab rig. Waste also named speed. One dab could feel like several hits from foil, but people seldom stopped at one, and with bigger hits, more of them, closer together (Section 3.2.3), a supply went much faster than on foil. Dabbing was not frugal, yet people without money to spare had to make a supply last; in November 2026, one participant described balancing the two: “I take small taps also very lightly to conserve and I actually rotate between dabbing and the oil burner to conserve since the stuff is so expensive and dabbing goes through it faster.” Alex and Jane disagreed about which waste counted:

Field note, April 2026: When Jane finishes, she sets her dabber down on a silicone mat, still emitting smoke. Alex is horrified — “You’re wasting it!” — and picks it up, inhaling the smoke still emanating from it. Jane is unmoved: “It’s a lot of time that you waste trying to get every last drop. Not like it makes a difference anyway. It’s never enough.”

Nearly everyone we observed valued the residue (“reclaim”), even those whose money made it unlikely they would ever need to smoke theirs. Residue built up inside ceramic tips, sometimes narrowing or blocking the opening, and it was often stored, a reserve against withdrawal that one participant likened to a fentanyl savings bank, which made empty dabbers targets of theft too. People had various ways of getting at the reclaim in dab rigs and dab-mounted bongs; one evaporated the water from his dab bong and smoked the gummy residue left behind (Fig. 6b). “I know I’m gutter as f***,” he joked, remarking on the foul taste, “but I can’t just throw drugs away. That’s drug abuse!”

Fine-grained knowledge of materials, temperatures, doses, and bodily sensation marks competence and belonging, consistent with the early signs of fentanyl connoisseurship described in San Francisco (Mars et al., 2024). Hierarchies of perceived risk attach to methods: people who dab are often seen within this community as less likely to overdose than those who smoke on foil, a view people who dab endorsed themselves (“I wouldn’t want somebody who only smokes on foil to take a fat rip off my dabber”), and the distinction extends to sellers, who read method as a tolerance signal:

“Some of those plugs won’t even sell to you. If you’re still on [foil], you’re seen as an overdose risk — especially if you’re trying to cold cop and buy from strangers, and they’re worried that you’re gonna die and bring them heat. But if you whip out your dabber and take a fat dab, they’re like, ‘Oh, shit, look at that.’ It proves that your tolerance is pretty high.”

This perception ran in one direction: people who smoked only on foil did not describe dabbing as riskier or their own hits as smaller, having no basis for comparison without dabbing themselves, and those who did compare rarely returned to identifying as “foil smokers.” Participants described how dabbing both escalates tolerance and becomes a visual marker of it to others. Participants also recognized tolerance losses after breaks: “Even if you take a few days off, you can noticeably feel your tolerance go down.” When people returned to use after incarceration or a period of abstinence, others advised them not to dab.

Stylistic variation itself carried social meaning. For Alex, dabbing is a ritual he visibly savors — the assembly, the heat management, the hardware — and his pride in his equipment is a form of expertise display. Jane “dabs to get high”: a plain silicone dab straw with a ceramic nail, “no frills, nothing special…reliable, cheap.” She rolls her eyes when he shows off a new build and hits her simple dab straw again. The contrast — technique and equipment as identity for some, bare delivery system for others — parallels the stratified connoisseurship described among people smoking fentanyl in San Francisco (Mars et al., 2024).

#### 3.2.6. Burns, fires, and user-generated mitigation

Burns and fires were a recurring concern among participants, which has also been noted in recent clinical data from the West Coast (Englander et al., 2026); one participant observed that “everybody I know that dabs has burnt themselves.” Even a brief loss of consciousness while holding a lit butane torch can ignite a fire. The most commonly reported burns came from the still-hot tip contacting skin or objects: participants described “perfect circular burns from the tip” on hands, arms, and legs (Fig. 9), and dabbers that had melted bedding and car seats. Torch burns were considered less dangerous than tip burns because the flame’s heat is felt approaching the skin (“as soon as you get close, it wakes you up”), the illusion of control, while a hot ceramic nail was seen as striking silently, almost at random. Fentanyl residue within the tip also has a tendency to liquefy and drip out; participants described this liquid as “lava,” and it caused severe burns when it dripped onto skin, instantly cooling and solidifying — “like napalm getting on you,” lamented a participant who described having to pull off a stiff, candle wax-like dollop of residue, watching the first couple layers of their skin come up with it, and then having to decide whether or not the resulting debris should be smoked or thrown away.

**Fig. 9.**
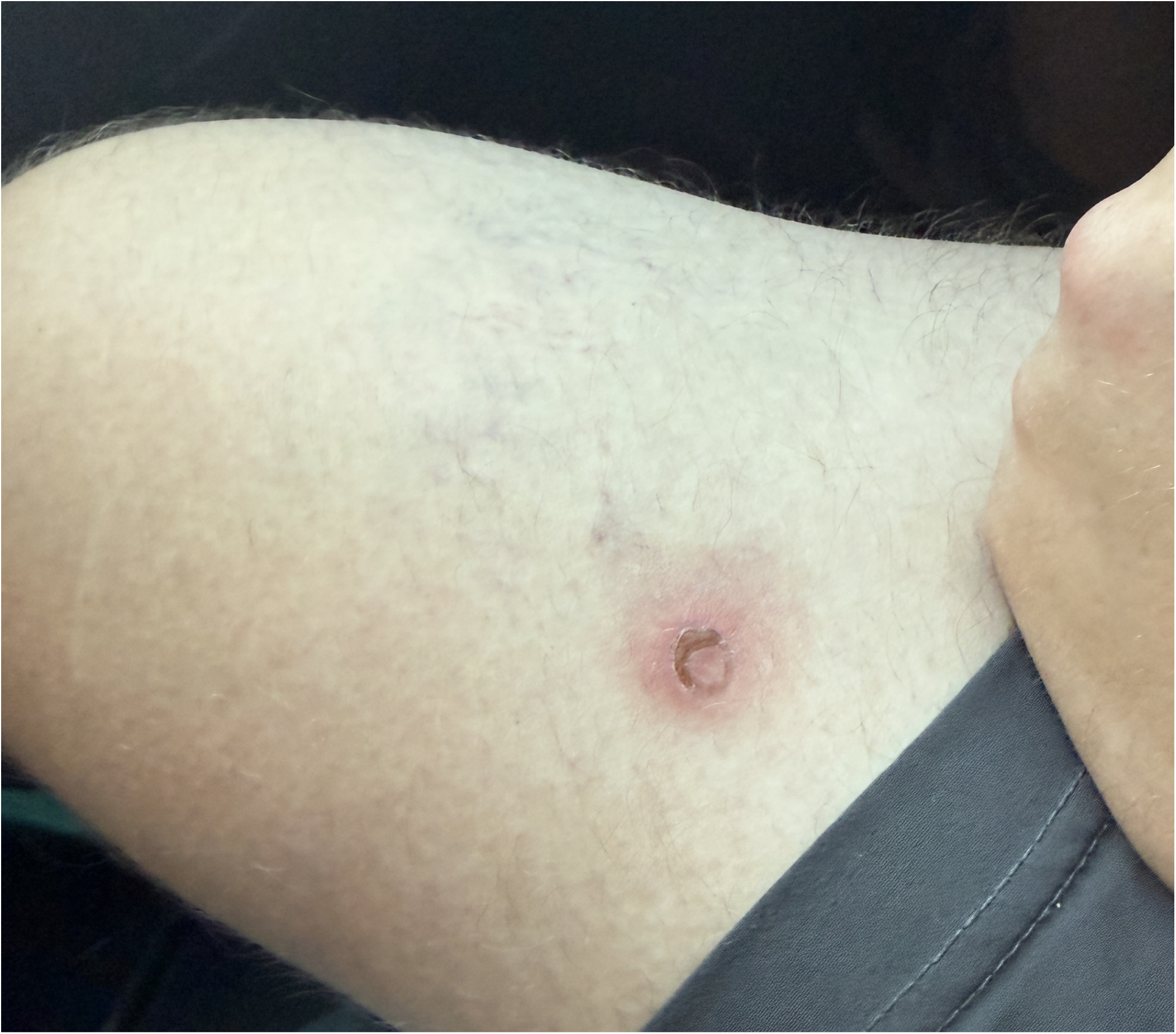
Contact burn from accidentally touching a hot ceramic dab nail to his thigh, about three weeks after injury, the most common if least severe burn pattern observed.

“This was late summer of 2024. I’m in shorts, and because the dope’s so shitty, you’re just firing through it — don’t care about waste. I took a hit and went to put the dabber down, and this hot lava — melted fetty — dripped out the back of my dabber onto my bare thigh. And, like, instantly cooled and solidified. It burned the shit out of me…A half hour later, it happened again. A half hour later, it happened again. So I had these three quarter-sized wicked burns, and they just would not heal. And people were like, ‘Oh, how’d you burn yourself?’ And I’m like, ‘Oh — hot oil, on the stove.’”

— Chris

The torches themselves failed in hazardous ways, the smallest and cheapest most of all when overused. We observed balls of flame engulfing the lighter, contact points hot enough to burn the hand holding it, and torches that kept emitting gas after the trigger was released, so the valve had to be closed to stop the flame. The flint often went first: the torch would hiss butane but not catch. One participant called the fix “jump starting.” With two hands, he pressed the trigger on the dead torch and brought a second, working torch to it, angling both flames so as not to burn himself, often a $4 gas-station torch relighting a far more powerful one. Cars, full of plastic and fabric, had nowhere to set a hot torch, and any makeshift surface shifts when the car moves. Startled by a passer-by, Jane set her still-lit torch on her car seat to pick up her nicotine vape and look like anyone else. It burned deep into the upholstery, now hidden under a seat cover that is itself riddled with burns.

Mitigation strategies were user-generated and borrowed, like the equipment itself, from cannabis dabbing. Chris no longer dabs in shorts and never passes the dabber over his legs, laying inexpensive, paper-thin silicone mats “protectively on everything” — “there’s products made for this, or, I don’t know, maybe they’re made for us” — and others used silicone pouches to store hot equipment between hits. Participants also rejected specific equipment as too dangerous: “Some of the monster torches have flame locks, but that just seems like a recipe for disaster — you could nod out and drop it on the ground and it would still be flaming, just setting everything on fire.” That participant was observed nodding out while torching their nail for another hit, bringing the still-flaming torch across the back of their hand. One participant also worried about the exposure inherent to heavy torch use: “I’m burning through so much butane — that can’t be good for my lungs.”

## 4. Discussion

In our Los Angeles sample, 87% of participants who reported a smoking method reported dabbing at least once, and dabbing exceeded foil as the most commonly reported method. Although we report dabbing as a smoking method to stay consistent with the survey, it is a discrete, novel route of fentanyl administration: the drug is vaporized off a separately heated intermediary rather than heated directly by flame, and it comes with its own equipment, technique, and patterns of dosing. We pair this with a sustained ethnographic inquiry that situates this finding within the broader transition from injection to smoking, with its own material culture, social meanings, and risks. Ethnographically, we observed most participants moving toward dabbing during our fieldwork. Similarly, in survey data, all 11 participants who initially reported another smoking method and returned on a later date subsequently reported dabbing (Table 2). This supports the ethnographic accounts while leaving the timing of adoption uncertain. Participants’ descriptions of an injection-like “rush” raise a pharmacokinetic hypothesis, informed by a clinical study of a controlled fentanyl aerosol rather than street dabbing.

In an open-label crossover trial, inhaled fentanyl vapor reached the blood as completely and as fast as an injection (MacLeod et al., 2012): ten healthy volunteers received 25 µg of fentanyl as a thermally generated aerosol, flash-heated off metal foil and taken in one brisk, complete inhalation with the breath held, and on another day the same dose intravenously. Absolute bioavailability, relative to the dose the device emitted, was 96.8%; peak arterial concentration and total exposure were similar to injection; and the slightly earlier inhaled peak was attributed to circulation time from the arm vein to the arterial catheter. If illicit fentanyl behaved like the trial aerosol when dabbed, injecting would add neither speed nor size of effect. People who use fentanyl may have found a way to optimize the drug’s inherent pharmacology, and dabbing may be unusually well suited to approximating the trial’s conditions: the drug flashes to vapor on a pre-heated surface, travels a closed path to the mouth, and is taken in one hard, deep inhalation with the breath held.

The hypothesis is untested in people who use drugs, and the trial was small, funded by the device’s maker, and run in healthy non-smokers. A dab rig diverges from it at every point that can be measured. Street fentanyl is a mixture of uncertain salt form; tip temperature is uncontrolled; particle size is unmeasured; and drug is visibly lost, in smoke wisping out of a bong and in the thick cloud exhaled after a big hit, where the trial lost 3%. In a separate study comparing inhaled and intravenous pharmaceutical heroin, inhalation bioavailability was estimated at 52% (Rook et al., 2006). This is a comparison within heroin, not of fentanyl dabbing with foil. Purpose-built devices that heat fentanyl to a controlled temperature can vaporize it with little degradation (Vazda et al., 2021), and a dab rig’s heated intermediary keeps the drug away from a flame of variable temperature. Letting a glowing tip cool before use fits fentanyl’s thermal chemistry: visible red heat begins at roughly 525 °C (Draper, 1847), whereas fentanyl has been reported stable to about 350 °C under flash pyrolysis, with degradation products by 500 °C (Manral et al., 2009). Cannabis e-nail users posting online cite a preferred temperature of about 378 °C (Meehan-Atrash et al., 2017). How fast a torch-heated tip cools has not been measured, and the hydrochloride salt, common in illicit fentanyl and far less volatile than the free base at ambient temperature (Ewing et al., 2022), has not been characterized at dabbing temperatures; for heroin, salt or base makes a large difference to how efficiently the drug volatilizes (Huizer, 1987). Whether dabbing delivers more fentanyl than foil is unmeasured. However, the perceived difference is large enough to organize individual preference and market behavior.

Most of what is known about dabbing comes from cannabis research, where concentrate use has been associated with higher THC exposure (Bidwell et al., 2020), stronger self-rated negative effects (Chan et al., 2017), and greater tolerance and withdrawal (Loflin & Earleywine, 2014) than smoked flower, and more frequent use has been associated with physical dependence (Meier, 2017). The analogy is imperfect: people who dab fentanyl use the same drug product as those on foil. Still, participants broadly reported single inhalations with a volume of vapor far exceeding that which is possible when smoking on foil, with implications for overdose risk and tolerance. What happens to adulterants and bulking agents such as xylazine, medetomidine, BTMPS, and lidocaine (Shover, Godvin, et al., 2025; Shover, Koncsol, et al., 2025) at dabbing temperatures is unknown. Among our samples, xylazine in samples sold as fentanyl was detected in 23% of samples (82 of 349) in 2024, and its median concentration was 0.1% among 106 quantified samples.

The high tolerance that dabbing likely indicates may itself be protective against opioid overdose. Two measured quantities suggest its scale: mean reported daily consumption of 1.07 g of drug product among 47 respondents in our previous Los Angeles study (Godvin et al., 2026) and mean fentanyl purity of 16.3% among 128 intact powders quantified between October 2025 and September 2026 (Drug Checking Los Angeles, unpublished data) together imply roughly 175 mg of fentanyl loaded per day, with an uncertain amount delivered. The people we observed from 2024 to 2026 were, after all, the survivors, more than a decade into an overdose crisis that has taken the lives of hundreds of thousands of Americans and multiple friends of every single person we talked to.

Dabbing is most likely to negatively impact the organs of the lungs and the skin. Acutely, the most salient harm is opioid-induced respiratory depression, and dabbing’s large, quickly inhaled boluses of vapor could shorten the interval between onset and apnea; sedation also brings a risk of aspiration, and aspiration pneumonia was diagnosed in 11% of hospital admissions for accidental opioid overdose in British Columbia (Morrow et al., 2019). The pulmonary consequences of smoking fentanyl chronically have not been characterized. Heroin smoking has been associated with pneumonia among HIV-negative drug users (Mientjes et al., 1996), and chronic obstructive pulmonary disease has been reported among people who smoke heroin (Bitar et al., 2021; Nightingale et al., 2020). The hardware adds its own exposures: butane gas, open flame, carbon monoxide, super heating and then inhaling off of material not manufactured to withstand those temperatures. Of particular concern were the cheap metal tips that arrived pre-installed in Amazon dabbers of dubious supply origin, always sold for some other ostensible purpose and repeatedly reported to leave a strong metallic taste when heated, making multiple participants question how they could possibly be safe for their intended purpose.

Dabbing’s capacity for rapid redosing of high-volume, large hits may raise overdose risk relative to foil. Going through fentanyl faster may also mean running out sooner and buying again more often, and because participants felt their tolerance decline after even a few days off (Section 3.2.5), a return to large hits after a gap may outpace the tolerance that remains. This is compatible with tolerance being protective: tolerance protects the person who has it, and the danger concentrates in the person whose tolerance has fallen, after jail, detox, or a hospital stay, while the equipment and the size of the hit stayed the same. At the same time, participants who had injected for years described dabbing as the thing that finally replaced it, and if that holds more widely, dabbing could help some people inject less often, and so suffer fewer invasive bacterial infections (Islam et al., 2019), skin and soft-tissue infections (Megerian et al., 2024), and other injection-associated harms (Karandinos et al., 2026). The evidence for substitution is ethnographic, including accounts from participants accustomed to injecting on the East Coast; the survey cannot establish whether dabbing replaced injection, and the two coexist: 5 of 8 participants who reported injection also reported dabbing. Dabbing may reduce injection, replace it temporarily when veins fail, or offer an alternative to returning to it. Opioid injection has declined among people who inject drugs in San Diego (Eger et al., 2024) and smoking has risen sharply in the South (Chen et al., 2024); what drives such shifts, and whether dabbing could contribute to them, warrants evaluation.

Englander and colleagues (2026) found acute-care burn encounter rates 4.3 times higher among Oregon Medicaid members with a diagnosis code for smokable drugs. In qualitative interviews, butane torch use appeared be the primary vector of burn risk, although the Medicaid data did not specify route of use and dabbing was not examined separately. (Butane torches are sometimes also used to smoke methamphetamine out of pipes, their use is not exclusive to dabbing nor opioids.) Our participants’ confidence that a torch cannot burn a sleeping person for more than a second or two, because the heat “wakes you up,” is contradicted by that work: people interviewed in Oregon described burns from passing out with a torch locked on, and one recalled a friend who “burned right through his hand” without waking (Englander et al., 2026). Pairing opioid sedation with an open flame and a heated element almost certainly amplifies this risk.

Widespread dabbing complicates standard safer-smoking guidance. The usual kit of foil, tooters, or hammer pipes (Canadian AIDS Treatment Information Exchange [CATIE], 2025; Chung et al., 2025; Kielhold et al., 2025; Megerian et al., 2024) is of less use to someone who dabs. Programs encountering dabbing should warn that one inhalation can deliver a much larger dose than people used to foil expect; caution against flame locks and the most powerful torches; and share heat-resistant surfaces and tools already used by people who dab.

### 4.1. Limitations

The survey sample was people who chose to use drug checking services in Los Angeles County; it may not generalize to all people who use fentanyl, locally or elsewhere. We have no ROA prevalence data for the other West Coast cities where we did fieldwork. Tools and lexicon looked much the same in every city observed, though we cannot say whether dabbing was equally common in each, and the sameness may partly reflect snowball recruitment through interconnected networks. The ethnography also has little to say about pipes, which the survey shows were used nearly as often as foil. Many of the participants most prevalent in our fieldwork were housed, at least marginally; we also observed dabbing among unhoused people, but the present analysis did not assess differences by housing status. ROA was reported for the sample tested, not for all of a person’s use. Geographic variation in fentanyl markets limits transferability, particularly outside the western United States where smoking is less common (Friedman & Shover, 2023; Karandinos et al., 2024; Shover et al., 2020). The smoking-method survey item followed from ethnographic observation, so the two data strands are not independent. No systematic health-outcome data were collected, and the health implications discussed here are hypotheses. Triangulating long-term observation with self-report and quantitative drug checking data narrows these constraints without removing them (Friedman, Montero, et al., 2022).

The smoking-method question was introduced after the route-of-administration question and was not consistently completed, particularly early in implementation, so more encounters have route responses than answers about smoking method. Missing smoking-method answers do not indicate absence of dabbing, and prevalence among respondents with a recorded method may differ from prevalence among all smokers. First-answer comparisons cannot establish when participants first adopted dabbing. Free-text answers naming a bong or a crack pipe were coded as dabbing (Section 2.3), which is why Table 1 records no other identifiable smoking method. Repeated encounters may reflect multiple samples submitted on the same date so we therefore restricted our analysis to encounters occurring on different days.

## 5. Conclusion

Dabbing predominated in this sample, yet remains largely unstudied. Participants recognized the gap: “Nobody’s talking about it. Everyone I know is doing it. We’ve been doing it for years now…They don’t even know it exists.” Research should measure dose delivery relative to foil, bioavailability, pulmonary effects, adulterant behavior when heated in this fashion, and the frequency and severity of burns. Dabbing’s potential to replace injection or avert a return to it warrants investigation alongside its risks. These questions require evidence-based, user-informed harm reduction responses that distinguish among smoking methods.

## Declarations

CRediT authorship contribution statement. Morgan Godvin: Conceptualization, Methodology, Investigation, Formal analysis, Visualization, Writing – original draft, Writing – review & editing. Levi Cecil: Investigation, Resources, Writing – review & editing. Adam Koncsol: Investigation, Formal analysis, Data curation, Writing – review & editing. Caitlin A. Molina: Investigation, Writing – review & editing. Megan Grabill: Investigation, Writing – review & editing. Ruby Romero: Investigation, Writing – review & editing. Elham Jalayer, Naomi Villanueva, Oscar Arellano, Amanda Cowan: Resources (community partner organizations hosting drug checking), Writing – review & editing. Russell G. Buhr: Writing – review & editing (pulmonary expertise). Honora Englander: Conceptualization (burn injury), Writing – review & editing. Joseph R. Friedman: Methodology, Writing – review & editing. Philippe Bourgois: Conceptualization, Methodology, Supervision, Writing – review & editing. Chelsea L. Shover: Investigation, Supervision, Project administration, Resources, Writing – review & editing.

## Ethics approval and consent

The ethnographic work was approved by the University of California, Los Angeles Institutional Review Board (IRB-21-0901). The quantitative and drug checking methodologies were approved by the same board (IRB-22-0760 and IRB-22-1273), and some aspects of the work were considered exempt from review as public health surveillance. Participants gave consent for interviews, audio recording and photography, including publication of the photographs in Figs. 6b and 9.

## Funding

Support for this research came from the Centers for Disease Control and Prevention’s Overdose Data to Action: LOCAL program (CDC-RFA-CE-23-0003).

## Declaration of competing interest

The authors declare that they have no competing financial or non-financial interests.

## Data availability

The drug checking data are not publicly available, to protect participant confidentiality.

## Declaration of generative AI use

During the preparation of this work the authors used Claude (Anthropic) and Codex (OpenAI) to edit and organize drafts, check the accuracy of references, rerun descriptive statistics, and prepare submission materials. Claude also drew the explanatory diagrams (Figs. 2 and 3) from basic shapes under the authors’ direction and cropped, rotated, and resized photographs for figure layout; no generative image model was used, and no photograph was generated or altered in content. After using this tool, the authors reviewed and edited the content as needed and take full responsibility for the content of the publication.

## Data Availability

Drug checking data about individuals is not publicly available in order to protect confidentiality, though drug checking results data are posted online.

https://www.drugcheckinglosangeles.com

## Notes

### Competing Interest Statement

The authors have declared no competing interest.

### Author Declarations

The Institutional Review Board of the University of California, Los Angeles approved the ethnographic work (IRB-21-0901) and the quantitative and drug checking methodologies (IRB-22-0760 and IRB-22-1273). Some aspects of the work were considered exempt from review as public health surveillance.

